# Modeling reductions in SARS-CoV-2 transmission and hospital burden achieved by prioritizing testing using a clinical prediction rule

**DOI:** 10.1101/2020.07.07.20148510

**Authors:** Jody R. Reimer, Sharia M. Ahmed, Benjamin Brintz, Rashmee U. Shah, Lindsay T. Keegan, Matthew J. Ferrari, Daniel T. Leung

## Abstract

Prompt identification of cases is critical for slowing the spread of COVID-19. However, many areas have faced diagnostic testing shortages, requiring difficult decisions to be made regarding who receives a test, without knowing the implications of those decisions on population-level transmission dynamics. Clinical prediction rules (CPRs) are commonly used tools to guide clinical decisions. We used data from electronic health records to develop a parsimonious 5-variable CPR to identify those who are most likely to test positive, and found that its application to prioritize testing increases the proportion of those testing positive in settings of limited testing capacity. To consider the implications of these gains in daily case detection on the population level, we incorporated testing using the CPR into a compartmentalized disease transmission model. We found that prioritized testing led to a delayed and lowered infection peak (i.e. “flattens the curve”), with the greatest impact at lower values of the effective reproductive number (such as with concurrent social distancing measures), and when higher proportions of infectious persons seek testing. Additionally, prioritized testing resulted in reductions in overall infections as well as hospital and intensive care unit (ICU) burden. In conclusion, we present a novel approach to evidence-based allocation of limited diagnostic capacity, to achieve public health goals for COVID-19.

**One Sentence Summary:** A clinical prediction rule to prioritize SARS-CoV-2 testing improves daily case detection, flattens and delays the curve, and reduces hospital burden.

## Introduction

Coronavirus disease 2019 (COVID-19) has caused over 10 million cases and 500,000 deaths globally as of July 1, 2020 (*1*). Rapid identification of cases is critical for managing an epidemic, as it allows for case identification, isolation, and contact tracing of infectious people, reducing transmission. However, many countries, including the United States (US), have experienced shortages in diagnostic testing capacity (e.g. test kits, swabs, etc.), and these shortages will likely continue in many settings worldwide (*2–4*). With limited test availability, decisions regarding who to test are often left to clinicians or health systems, with potential guidance from national and international health authorities. The rationing of SARS-CoV-2 (the virus responsible for COVID-19) testing may default to those with more severe disease or at higher risks of complications, as definitive diagnosis is most critical to guide care for such patients. However, because of their symptoms, severely ill patients may also be more likely to self-limit contacts, thereby limiting the indirect benefit of their diagnostic testing on reducing transmission.

When diagnostic testing is unavailable, clinical case definitions are used instead in clinical management and public health response (*5*). From its emergence in humans in late 2019 until early April 2020, the Centers for Disease Control and Prevention (CDC) defined a clinical COVID-19 case as having cough, shortness of breath, or fever. Despite recent refinements in the case definition (*5, 6*), and the identification of olfactory and gustatory dysfunction to be highly specific (*5, 7, 8*), the majority of symptomatic persons with SARS-CoV-2 infection present with symptoms that overlap with those of other common respiratory infections, including respiratory syncytial virus and influenza (*9*). Difficulties in differentiating between infectious etiologies based on clinical presentation can cause delays in diagnosis and treatment of individual patients, as well as in public health responses that curb transmission (e.g. contact tracing).

Given limited testing capacity and the non-specific symptom profile of COVID-19 disease, tools to guide clinicians in how and when to use limited testing capacity are needed. Clinical prediction rules (CPRs) are commonly used tools to help to guide clinical management decisions, such as who should undergo testing or receive limited clinical resources. They provide standardization and consistency in care between physicians, as well as improved diagnostic accuracy (*10*). Some widely used CPRs include the Centor criteria (*11*) for diagnosis and treatment of strep pharyngitis, the Ottawa ankle rule (*12*) for appropriate use of X-ray in setting of ankle trauma, and the CURB65 score (*13*) for triage of patients with pneumonia. As CPRs are usually developed to improve patient care, their evaluation has been focused on their impact on patient-level outcomes; the impact of CPRs on population health, including on transmission dynamics of infectious pathogens, has not been widely studied.

Compartmental models (e.g. SEIR or “susceptible-exposed-infected-removed” models) are often used to describe disease dynamics through a population. They combine epidemiological information (e.g. transmissibility, duration of infectiousness, reproductive number) to provide a picture of the population-level disease dynamics over time (*14, 15*), but have not yet been used to evaluate the impact of CPRs on population-level public health outcomes. We use an SEIR model of SARS-CoV-2 transmission to evaluate the population-level impacts of a clinical prediction rule for prioritization of testing in a setting where tests (or testing supplies) are limited. We define prioritized testing as the allocation of SARS-CoV-2 tests to patients more likely to be infected, as predicted by a CPR. Within the SEIR model, those who test positive are immediately removed from the infectious class (i.e. are isolated and stop transmitting disease), lowering the realized transmission rate.

Our primary objective was to measure potential impacts of prioritized testing (using the CPR) on the course of the SARS-CoV-2 pandemic, including the magnitude and timing of the outbreak peak as well as the associated impact on hospitalization and intensive care unit (ICU) burden.

Additionally, we determined the conditions (e.g. test availability, test seeking volume, effective reproductive number) in which prioritized testing may result in the greatest reduction of SARS-CoV-2 infections and hospitalizations.

## Results

### A parsimonious 5-variable clinical prediction rule identifies those who would test positive for SARS-CoV-2

To develop a clinical prediction rule to prioritize SARS-CoV-2 testing based on probability of testing positive, we used data gathered from electronic medical records of patients tested for SARS-CoV-2. Data were gathered from a period where testing eligibility was based on presenting with at least one of cough, fever, shortness of breath, or a high risk of exposure given recent travel or contact with a laboratory-confirmed case (March 1, 2020 – April 6, 2020). We use the phrase *test eligible* to describe any person seeking a test who satisfies these conditions. We used a logistic regression to develop a parsimonious 5-variable CPR which included age, Area Deprivation Index (ADI) (*16*), prior exposure, smoking status, and history of travel. Our CPR had a cross-validated AUC of 0.69 (95% CI: 0.68 - 0.70). We explored using additional variables but found this only marginally improved predictive ability (AUC up to 0.71; Fig. S1 and Table S1), at the expense of requiring much greater data entry effort by clinicians. We also considered alternative versions of the CPR in light of the varying availability of predictor variables in different clinical contexts. We explored models excluding symptoms, including vital signs, and including a race/ethnicity variable. Again, these did not meaningfully improve predictive ability (AUC up to 0.72; Table S1). Finally, we explored using random forest regression to fit the models, but logistic regression estimates had consistently higher AUCs.

### Prioritized testing increases the proportion of those testing positive in settings of limited testing capacity

In settings of limited testing capacity, we found that a prioritized testing approach using a CPR resulted in testing a greater proportion of those infected with COVID-19 compared to indiscriminate testing (Fig. 1). When comparing indiscriminate testing to prioritized testing, the absolute difference in the number of people infected with COVID-19 who were tested was greatest for intermediate levels of testing availability, achieving the greatest benefit to disease detection when between 40-60% of test eligible people received testing (vertical difference between solid lines in Fig. 2). However, the proportional increase in the number of people infected with COVID-19 who were tested was greatest for low testing capacity, with the largest fold changes seen when <20% of test eligible people received testing (dotted line in Fig. 2). For example, if the rate of SARS-CoV-2 positivity among test eligible people was 5% and there was test capacity for only 10% of those test eligible people, we would expect to see a nearly 3-fold increase in the number of patients testing positive on a given day if using prioritized testing instead of indiscriminate testing (Fig. 2A).

**Fig 1.**
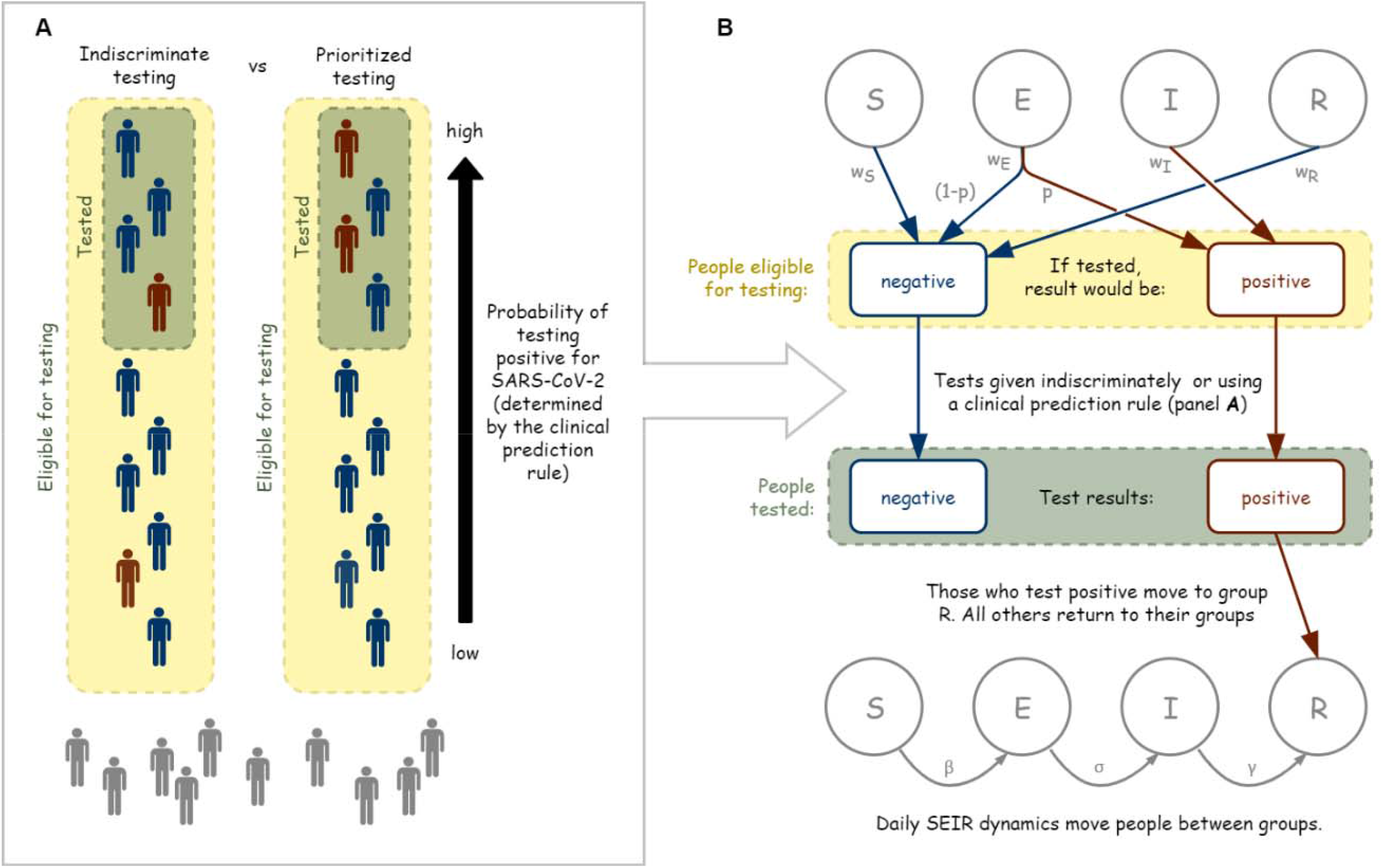
Effects of prioritized testing on daily testing outcomes and incorporation into an SEIR model. **(A)** Schematic comparing the testing of a subset of test eligible people using either indiscriminate testing or prioritized testing. Red figures would test positive for SARS-CoV-2 and blue figures would test negative. Gray figures are not seeking tests. For prioritized testing, people are arranged and then tested according to their probability of testing positive, as determined by the clinical prediction rule. **(B)** Visual depiction of how prioritized testing was incorporated into the daily stochastic SEIR model. People in each compartment seek testing with probability *w*_*S*_, *w*_*E*_, *w*_*I*_, and *w*_*R*_. If tested, a proportion *p* of those in group *E* would test positive. Following testing, daily SEIR dynamics occur with transmission rate β, incubation rate σ, and removal rate γ.

**Fig 2.**
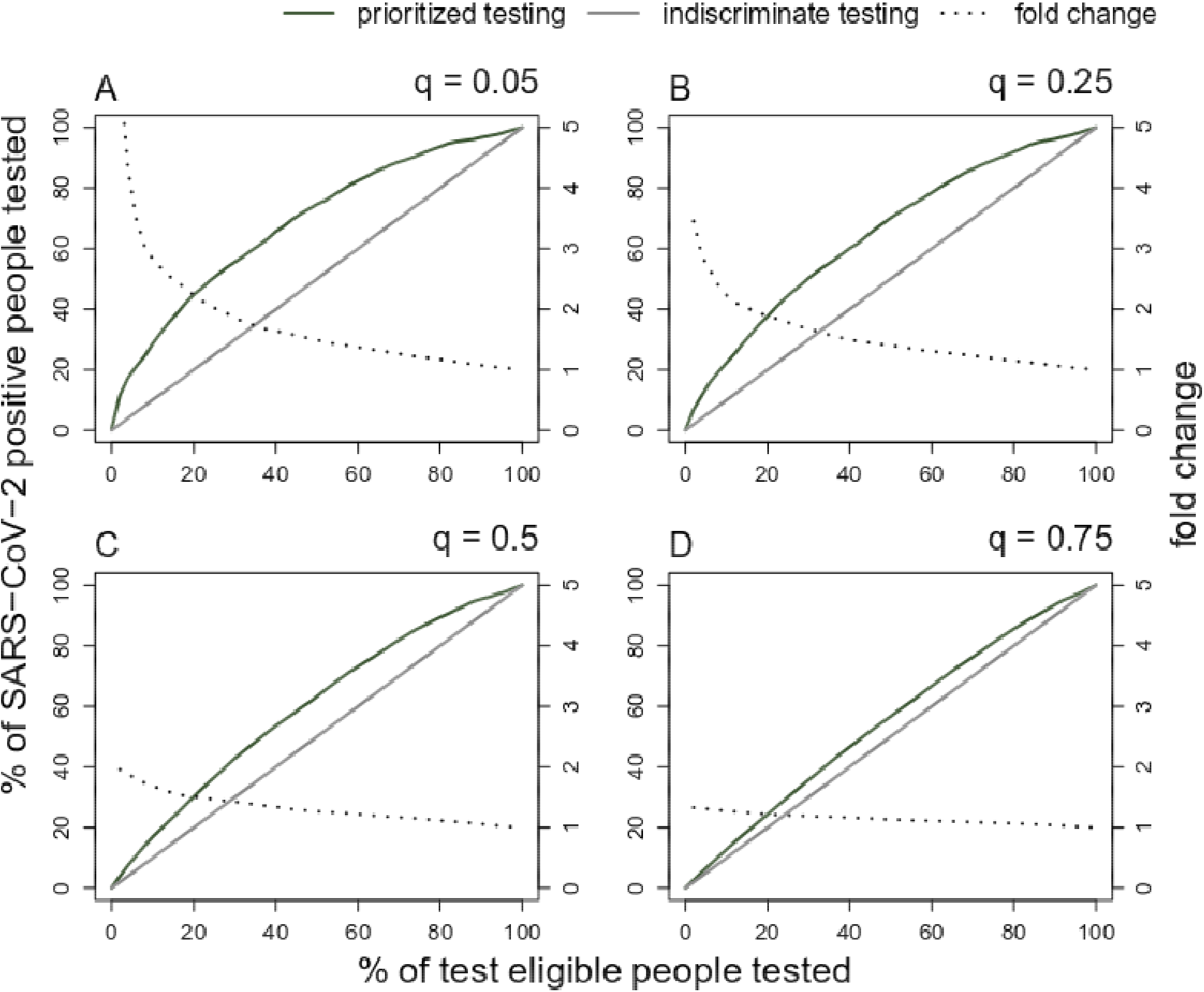
Effect of prioritized testing compared to indiscriminate testing on the percentage of SARS-CoV-2 positive, test eligible people, who are tested. The horizontal axis allows for comparison between different testing capacities. The vertical axis refers to the percent of SARS-CoV-2 positive, test eligible people. Dotted lines denote the fold change between the grey and green lines. The percent of SARS-CoV-2 positive people (proportion *q*) of those who are test eligible is 5% in **(A)**, 25% in **(B)**, 50% in **(C)**, and 75% in **(D)**.

These results were sensitive to the proportion of SARS-CoV-2 positive patients who are test eligible, with greater differences between prioritized and random testing strategies seen for low rates of COVID-19 positivity (compare Fig. 2A-2E). Results were robust to the total number of test eligible persons. The average empirical AUC values recorded during resampling were 0.68, consistent with the CPR model AUC.

### Prioritized testing delays and flattens the peak of infections

Using a stochastic SEIR compartmental model of transmission, we show that prioritized testing delays the timing and reduces the prevalence at the infection peak and reduces the total number of infections over the course of the pandemic (Fig. 3, Table 1). For our base parameter set, prioritized testing as compared with indiscriminate testing resulted in a 27 day delay in the timing of the infection peak and a 31% decrease in the peak number of infections.

**Fig 3.**
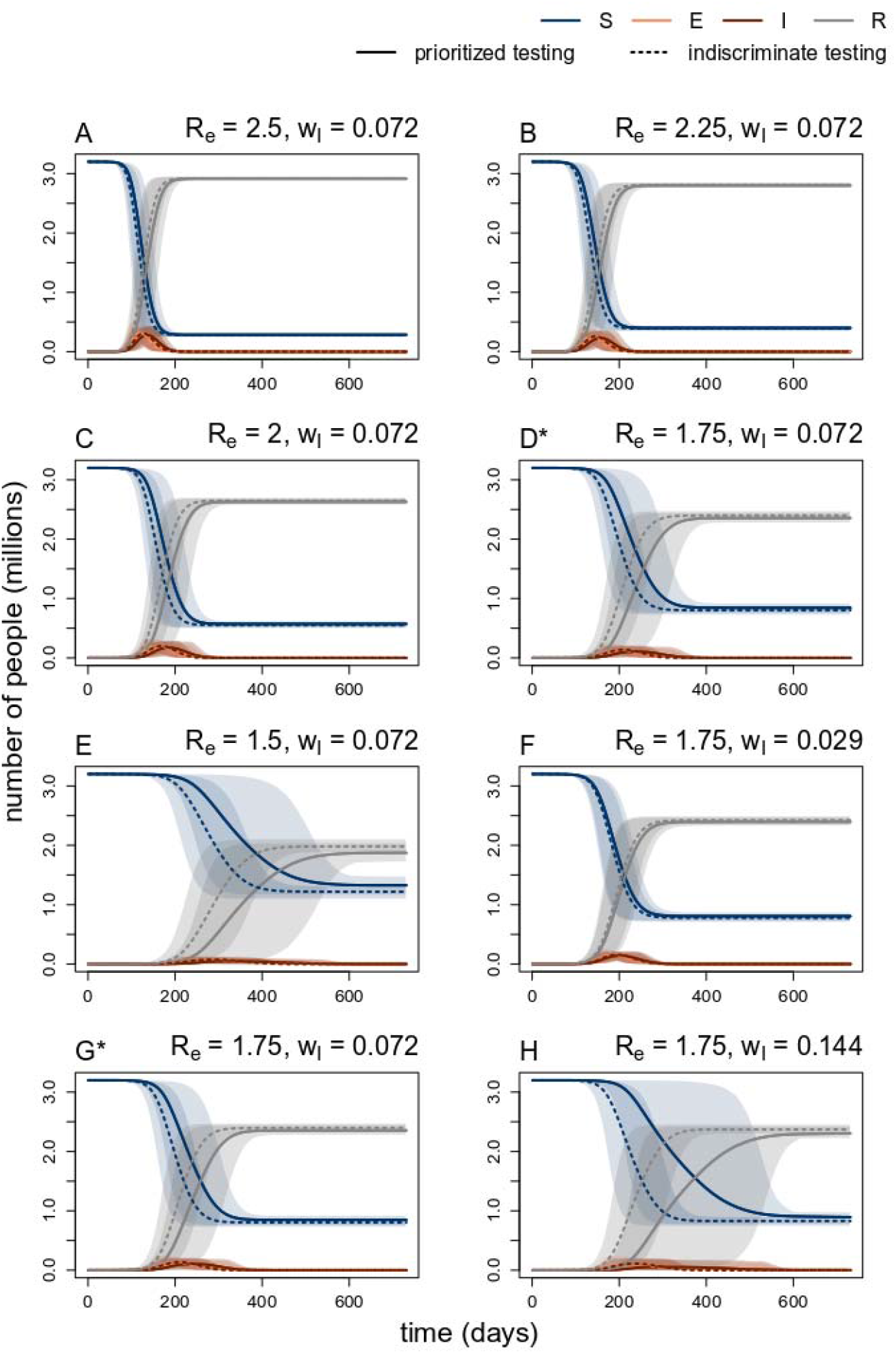
Comparison of SEIR curves between models with prioritized versus indiscriminate testing for decreasing values of the effective reproductive number, *R*_*e*_, **(A)-(E)**, and decreasing rates of test seeking among infectious individuals *w*_*I*_, **(F)-(H)**. Solid line are the mean of 1000 stochastic simulations with prioritized testing, and the dotted lines are the mean for the model with indiscriminate testing. Shaded regions denote corresponding middle 95^th^ percentiles of simulations. **(A)** *R*_*e*_ *=* 2.5, **(B)** *R*_*e*_ *=* 2.25, **(C)** *R*_*e*_ *=* 2.0, **(D)** *R*_*e*_ *=* 1.75, **(E)** *R*_*e*_ *=* 1.5, **(F)** *w*_*I*_ *=* 0.029, **(F)** *w*_*I*_ *=* 0072, **(F)** *w*_*I*_ *=* 0.144. *\**Note that **(D)** and **(G)** have the same parameters, but have both been included to show sequential change as we vary *R*_*e*_ and *w*_*I*_.

**Table 1.** Effects of prioritized testing on the SEIR infection dynamics over a range of parameter values. All parameter values are as stated in the text, except where stated otherwise in this table. Each column compares the mean results from 1000 stochastic simulations of the model with prioritized testing to one with indiscriminate testing. Bolded entries denote the results for the base parameter set described in the text (i.e., *R*_*e*_ =1.75, *w*_*I*_ = 0.072, *N*_*tests*_=1000), and are repeated for reference in each subsection. NA values exist where hospital or ICU demand did not exceed capacity for either the prioritized or indiscriminate testing model.

|  | Delay in peak timing (days) | Reduction in peak height (people) (%) | Reduction in total infections (people) | Reduction in people-days above hospital capacity (%) | Reduction in people-days above ICU capacity (%) |
| --- | --- | --- | --- | --- | --- |
| **$R_e$** |  |  |  |  |  |
| 2.5 | 8 | 28,114 (10%) | 6,610 (0%) | 29,550 (7%) | 145,706 (73%) |
| 2.25 | 12 | 19,494 (13%) | 10,469 (0%) | 38,842 (11%) | 139,152 (77%) |
| 2.0 | 18 | 27,344 (16%) | 20,224 (1%) | 50,091 (21%) | 126,428 (83%) |
| **1.75** | **27** | **32,321 (31%)** | **40,933 (2%)** | **76,401 (85%)** | **108,707 (98%)** |
| 1.5 | 36 | 26,441 (56%) | 108,342 (5%) | NA | 41,616 (100%) |
| **$w_I$** |  |  |  |  |  |
| 0.029 | 7 | 14,570 (10%) | 28,433 (1%) | 34,173 (26%) | 108,616 (89%) |
| **0.072** | **27** | **32,321 (31%)** | **40,933 (2%)** | **76,401 (85%)** | **108,707 (98%)** |
| 0.144 | 49 | 54,255 (97%) | 66,352 (3%) | 23,023 (100%) | 92,076 (100%) |
| **$N_{tests}$** |  |  |  |  |  |
| 500 | 17 | 28,588 (23%) | 20151 (1%) | 66,344 (50%) | 114,920 (93%) |
| **1000** | **27** | **32,321 (31%)** | **40,933 (2%)** | **76,401 (85%)** | **108,707 (98%)** |
| 3000 | 26 | 15,697 (33%) | 137,680 (6%) | NA | 31,098 (100%) |
| 5000 | 2 | 2,165 (8%) | 121,334 (7%) | NA | NA |

**Fig 4.**
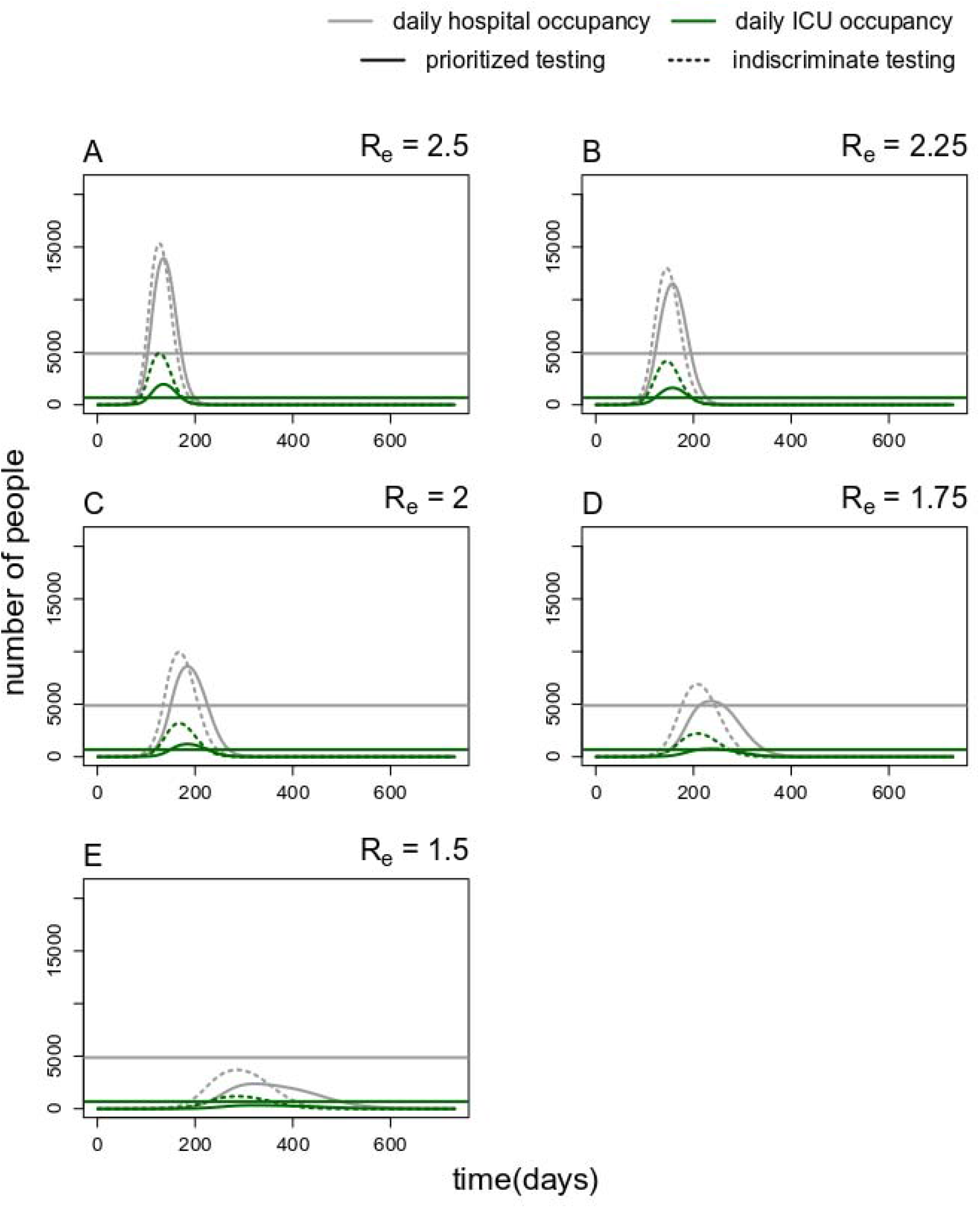
Comparison of simulated demand for daily hospital and ICU occupancy between models with prioritized versus indiscriminate testing. The solid line are the mean of 1000 stochastic simulations with prioritized testing, and the dotted lines are the mean for the model with indiscriminate testing. *R*_*e*_ decreases from 2.5 to 1.5 in increments of 0.25 in plots **(A)** through **(E)**.

The differences in the timing and numbers of infections between a model with prioritized versus indiscriminate testing were greatest for lower values of the effective reproductive number, *R*_*e*_ (Fig. 3, Table 1). *R*_*e*_ measures the average number of secondary infections from a single infected individual in an actual population with immunity, interventions, and any other factors that may impact disease spread. Note that here *R*_*e*_ refers to the effective reproductive number in the absence of testing, so the observed, time-varying effective reproductive number, *R*_*t*_, would be lower if a test-and-isolate strategy were also implemented.

Increasing the proportion of infectious test eligible people (*w*_*I*_) had a positive impact on the magnitude of the differences between the indiscriminate and prioritized testing models (Fig. 3, Table 1). Increasing the number of tests available (*N*_*tests*_) increased the differences for low values of *N*_*tests*_ but then had reduced benefits for higher values (Table 1), consistent with Fig. 2. When alternate CPRs with similar AUC values were considered, results varied only marginally (Table S1). Alternate CPRs with higher AUC values did not necessarily perform better on all metrics (Table S1). We also found a lack of sensitivity in results to the proportion of exposed individuals E who test positive for SARS-CoV-2 (*p*) (Table S1).

### Prioritized testing reduces hospital and ICU bed burden

Finally, we explored the impact of prioritized testing on hospital and ICU bed occupancy, basing our parameters on the outbreak in Utah. We demonstrated that prioritized testing resulted in reductions in the number of people-days (i.e., sum of the number of people on each day needing a hospital/ICU bed) where demand exceeded capacity for both hospital and ICU beds (Table 1). For our base parameter set, prioritized testing as compared with indiscriminate testing resulted in 85% and 98% reductions in the number of people-days above hospital and ICU capacity, respectively.

## Discussion

A clinical prediction rule to prioritize SARS-CoV-2 testing positively impacts both the number of laboratory-confirmed cases per day, as well as long-term disease dynamics when testing is scarce. We developed a CPR that predicts the probability that a given patient would test positive for SARS-CoV-2. These predicted probabilities can then be used to decide who receives laboratory testing. We incorporated this model of prioritized testing for our estimated AUC into our SEIR model and showed the value of our CPR, with appreciable delays in the timing and height of the infection peak, decreases in the total number of infections, and reductions in the number of people-days above hospital and ICU capacity. This novel combination of analytic methods allowed us to highlight both the individual- and population-level benefits of the CPR.

In spite of our CPR having only moderate discriminatory performance (AUC=0.69), our results show that prioritizing SARS-CoV-2 testing, even based on less-than-perfect CPRs, still has a meaningful impact on individual and population disease burden. Furthermore, future predictive models built following more extensive and improved data collection (e.g. standardized collection by clinicians over a longer time) may improve CPR performance, thereby further improving the impact of prioritized testing on community disease burden.

When considering the individual-level impact of the CPR on test-eligible individuals, we found that prioritized testing yielded the greatest absolute gains for intermediate testing capacity (capacity to test between 40-60% of test eligible people), and highest proportional gains for low testing capacity. Improved diagnostic triage through prioritized testing leads to diagnosis of individuals earlier in their course of disease, with potential for benefit through earlier initiation of therapies or medical monitoring, and isolation or contact-tracing precautions (*17*).

At the population level, we found notable impact of prioritized testing on COVID-19 dynamics, leading to reductions in overall infections and on hospital and ICU burden, as well as delaying the infection peak, providing more time for health systems to prepare for the surge. The magnitude of this impact was sensitive to several key parameters. For example, when *R*_*e*_ was lowered, as may happen with the introduction of other public health interventions such as social distancing, the effects of prioritized testing increased. This suggests that implementing prioritized testing concurrently with other non-pharmaceutical interventions that reduce *R*_*e*_, can help to maximize potential gains. Increasing the proportion of infectious people who seek testing (*w*_*I*_) increases the effects of prioritized testing because of the indirect benefit (reduction of *R*_*e*_) of isolating those individuals quickly. This may occur in populations with a higher proportion of symptomatic individuals, such as older populations (*18*) or those with other known risk factors (*19*). Alternatively, the proportion of infectious individuals seeking testing could be increased intentionally through interventions such as contact tracing or campaigns to encourage test-seeking behavior.

For any given level of testing, when COVID-19 is prevalent and comprises a large fraction of the test eligible population, either testing strategy can be impactful in reducing transmission by speeding up isolation. For any given level of testing, when COVID-19 is prevalent and a small fraction of the test eligible population, prioritized testing using the CPR leads to greater population level benefit. Thus, in settings with both COVID-19 and high prevalence of influenza-like illness (e.g., a possible fall and winter scenario), prioritized testing may be of increased value.

Use of prioritized testing is most useful in situations with limited test capacity, as the benefits of prioritized testing become negligible when test demand does not exceed test availability. While some health systems have increased their testing capacity to meet current demands, it is anticipated that demand will increase both as seasonal respiratory viral infections increase and in future waves of COVID-19. Further, many countries and regions with lower resources may continue to have limited capacity for testing. Investment in a system of prioritized testing may be more cost-effective than the manufacturing or purchasing of more tests to meet demand.

Additionally, this approach can be useful in future pandemic preparedness as a similar approach implemented in a timely manner may help maximize finite testing resources during the initial stages of a future outbreak, until adequate, affordable testing is available.

Our study has a number of limitations. Our CPR was derived using data from a single health system, with test eligibility criteria that followed CDC guidance from early in the pandemic; thus, as with other diagnostic CPR for SARS-CoV-2 (*20*), our CPR may not be generalizable and requires validation in other settings. Instead, we highlight the generalizability of the approach we have presented, and that the individual and population level impacts of prioritized testing are robust to the specific CPR used (Table S2). Secondly, our model assumes that all individuals seeking testing would present at the same time. In most clinical settings, the implementation of such a CPR would involve the use of a probability threshold, set based on data from the previous day(s) and the expected number of test eligible people. The optimal setting of this threshold, given stochastic testing demands and infection dynamics, would be an area for future exploration during clinical trials. Third, we did not consider the implications of the sensitivity and specificity of SARS-CoV-2 tests, as these values are not currently well known and evolving with new tests and optimization of sampling techniques. Low sensitivity and specificity in the diagnostic tests would reduce the utility of testing in general, and thus also of prioritized testing.

Our SEIR model assumes complete and immediate compliance of isolation by infectious individuals who test positive for SARS-CoV-2. While this is unlikely to be the case in reality, noncompliance with isolation is mathematically equivalent to reducing the number of tests available, which is included in our model. Conversely, if everyone with COVID-19 symptoms were to isolate perfectly, this would be equivalent to 100% testing of all eligible individuals and prioritized testing would have small marginal benefits. We note that testing is required for contact tracing, and more efficient confirmation of infected individuals would be expected to increase the efficacy of contact tracing (which we did not consider in our model). Finally, we assumed that infectious individuals who seek testing but do not receive a test have the same subsequent transmission behavior as those who do not seek testing. The transmission behavior of untested, infectious individuals includes other public health interventions such as social distancing or isolation if experiencing COVID-19 symptoms. We explored this possibility by varying the effective reproductive number, Re, which is reduced through other effective public health interventions.

To highlight key trends in a generalizable framework, we did not incorporate demographics such as age or socioeconomic status into the SEIR model. More sophisticated models designed for a specific population would yield more precise estimates of the effects of prioritized testing for that given population. Similarly, we have assumed interventions are constant and effect *R*_*e*_ consistently throughout each simulation. However, most interventions are transient, implemented and lifted in response to broader epidemiological, social, and economic cues. Models for a specific series of interventions would, again, provide more precise estimates of the effects of prioritized testing for that situation.

The limited availability of SARS-CoV-2 testing has hampered disease mitigation efforts in many locations. By incorporating a diagnostic clinical prediction rule into a transmission dynamics model, we have demonstrated the potential efficacy of prioritized testing for delaying and reducing peak infections and the consequent healthcare demand. By highlighting parameter regimes in which these effects are greatest, we have suggested situations in which it may be most efficacious to consider using a CPR to prioritize testing of testing shortages caused by the emergence of a novel infectious disease such as SARS-CoV-2.

## Materials and Methods

### Clinical prediction rule

#### Data

All patients tested for SARS-CoV-2 in the University of Utah Health (UHealth) system were eligible for our study. UHealth performed approximately 30% of all COVID testing in the state of Utah during our study period, March 1, 2020 thru April 6, 2020 (*21*). We created a near real-time electronic registry of all patients tested for SARS-CoV-2 at UHealth from the hospital operations dashboard. Testing for SARS-CoV-2 during the study period was predominantly nasopharyngeal swab samples using nucleic acid amplification methods. We combined this registry of tested patients with additional data on demographics, clinical symptoms, and patient characteristics from the Enterprise Data Warehouse (EDW) which aggregates data from disparate data sources within the UHealth system. Patients were entered into a RedCap registry for manual review, with the default sort by MRN. We conducted manual chart review of the clinical text collected 24 hours before and after SARS-CoV-2 testing. After about two weeks of reviewing, we selected patients to ensure we reviewed at least 20% of tested patients for each day through March 31, 2020. The remaining records were those completed by the time of these analyses. This resulted in 1,983 patient records. We collected data on symptoms and exposure risk factors. Symptoms/exposure factors not mentioned in the medical chart notes were extracted as ‘not mentioned.’ For analyses, we assumed any symptom not mentioned was not present. Additional observations were dropped due to missing data, for an analytic sample size of 1,928.

This study was reviewed by the University of Utah Institutional Review Board (IRB) and determined to be exempt.

#### Predictive variables

We considered the following variables in our clinical prediction rule: age (analyzed as a continuous variable); gender (female/male, too few patients reported non-binary gender to analyze separately); state ranked area deprivation index (*16*) based on home address zip code; smoking status (never, former, current, missing); binary variables for presence/absence of each of the following symptoms: cough, fever, shortness of breath, sore throat, nasal, headache, lethargy, myalgia, diarrhea, and nausea/vomiting; any comorbid condition; healthcare worker status (yes/no); history of out of state travel in the prior 2 weeks (yes/no); and exposure to a confirmed SARS-CoV-2 case (yes/no);

#### Statistical Analysis

We screened variables using random decision forests to identify the most predictive variables. Random forests are an ensemble learning method where multiple decision trees (1000 in this analysis) are built with only a random sample of potential predictors (sqrt of the number of parameters) considered at each split, thereby decorrelating the trees and reducing variability (*22*). Variables were ranked by predictive importance based on the reduction in variance achieved by including the variable in the predictive model. Generalizable model performance with the goal of discrimination was assessed using 5-fold cross-validation. In each of 100 iterations, random forest regression and logistic regression models were fit to a training dataset (random 80% sample of dataset) using the top *n* variables, where *n* ranged from 1 to all predictors described above. Each of these models were then used to predict the outcome (testing SARS-CoV-2 positive) on the test dataset, and the C-statistic (area under the receiver operating characteristic (ROC) curve (AUC)) was recorded. After examining the relationship between number of predictors used and model-performance (AUC), it was determined that the top 5 predictors in the logistic regression model would be used in the clinical prediction rule, as this balanced predictive ability with parsimony.

### Modelling the effects of prioritized testing on the proportion of those testing positive in settings of limited testing capacity

We first explored the effects of prioritized versus indiscriminate testing on the timescale of a single day (Fig. 1). On a given day, we assumed a certain number, *N*_*eligible*_, of people seek testing and are test eligible (have cough, fever, shortness of breath, or known exposure and seek testing). Of those, a certain proportion *q* would test positive for SARS-CoV-2 if given a test and the rest, (*1-q*), would test negative. We measured the proportion of test eligible, SARS-CoV-2 positive patients who received testing under the two testing regimes: prioritized and indiscriminate testing.

For each simulation, we sampled *N*_*eligible*_ people, with replacement, from the results generated during the 5-fold cross-validation of the clinical prediction rule, simulating the group of people eligible for testing that day. Each person in this group had both a known test result (positive or negative), as well as a probability of testing positive generated during cross validation of the clinical prediction rule described above. We also calculated the AUC of each of the simulated groups to check if it was consistent with the AUC of the clinical prediction rule.

We assumed a limited number, *N*_*tests*_, of SARS-CoV-2 tests were available daily. To model the indiscriminate testing scenario, we randomly sampled *N*_*tests*_ people for testing from the group of people eligible for testing. For the prioritized testing scenario, we ordered the list of test eligible people by their ascribed probabilities of testing positive for SARS-CoV-2 and tested the *N*_*tests*_ people with the highest probabilities.

Using the known SARS-CoV-2 test result for each person, we then checked how many people tested positive and compared this number between indiscriminate and prioritized testing strategies. We explored the effect of varying *N*_*eligible*_, the number of test eligible people (varied from 1000 to 50000), *q*, the proportion of test eligible people who were SARS-CoV-2 positive (varied from 1-99%), and *N*_*tests*_, the number of available tests, which is the same as varying the percent of test eligible people who are tested (varied from 0-100%). We simulated this scenario 1000 times for each set of parameters.

### Modelling the effects of prioritized testing on the peak and total numbers of infections

We also considered the effect of prioritized testing on disease spread in the population over longer time scales (months-to-years). We incorporated the same processes described above into a stochastic SEIR model parametrized for COVID-19. On each modelled day, we simulated the following steps (Fig. 1B): (1) Consider the number of individuals in each disease state—*S*, susceptible, *E*, exposed, *I*, infectious, and *R*, removed. Assume a fixed proportion of each group (*w*_*S*_, *w*_*E*_, *w*_*I*,_ *w*_*R*_) are have symptoms or a travel or contact history that would make them test eligible each day; (2) Assume that, if tested, those in groups *S* and *R* would test negative. A proportion, *p*, of those in group *E* would test positive, while (1-*p*) would test negative. Those in group *I* would test positive. (3) Of the test eligible group, test a subset based on the number of available daily tests, *N*_*tests*_. Select this tested subset either randomly or using prioritized testing with the CPR, as described above. (4) Assume those who test positive isolate immediately and effectively; move them into the *R* group. Those who test negative or are not tested remain in their original groups. (5) Apply the stochastic SEIR model (described below) to the population for one day. (6) Repeat steps 1-5 for a chosen number of days.

### Epidemiological model

We used a stochastic SEIR model (Fig. 1) to highlight the drivers determining the impact of prioritized testing on disease dynamics. We assumed frequency dependent transmission with rate β, incubation rate σ, and removal rate γ. We did not include any birth or deaths in the population and deaths from COVID-19 are included in the removed compartment. Stochasticity was introduced into the classical deterministic framework by letting the number of new exposed, infectious, and recovered cases be random variables, each drawn from a binomial distribution with the probability of “success’’ coming from the deterministic core model. All simulation code is archived and available online at doi:10.5281/zenodo.3924186.

Unless stated otherwise, parameters related to testing were *w*_*S*_ = 0.0013, *w*_*E*_ = 0.0013, *w*_*I*_ = 0.072, *w*_*R*_ = 0.00084, *p* = 0.7, and *N*_*tests*_ = 1000. SEIR parameters were σ = 1/5.2 (*23, 24*), γ = was pulled from a uniform distribution ranging from 1/7 to 1/4 (*25–28*), *R*_*e*_ = 1.75, and β = γ * *R*_*e*_ (*29*). For COVID-19, *R*_*0*_ is estimated between 2-3 (*23, 30, 31*) and we explored the effects of interventions (such as social distancing or contract tracing) implicitly by considering a range of effective reproductive numbers, *R*_*e*_, from 1.5 to 2.5. We also considered *w*_*I*_ values ranging from 0.029 through 0.144, and *N*_*tests*_ values of 500, 1000, 3000, and 5000. For full details on model parametrization, see *Parameterization* in the Supplementary Material.

We ran simulations assuming a total population of 3.2 million, the approximate population of the state of Utah (*32*). We assumed an initial condition of 15 people in the infectious class and all others in the susceptible class. We ran our simulations for a time period of 2 years. For each set of parameters considered, we ran 1000 stochastic simulations and then calculated the mean value of each of the *S, E, I*, and *R* classes as well as 95% prediction intervals.

We then calculated several metrics including the timing of the peak of the mean infection curve; the peak value of the mean infection curve; and the mean total number of infections by the end of the simulation. These metrics allowed us to compare expected outcomes between the models with indiscriminate testing and prioritized testing.

### Modelling the effects of prioritized testing on hospital and ICU burden

To highlight the associated implications for healthcare demand, we also modelled the daily occupancy of hospital beds and ICU beds by following the recommendations of the Centre for Disease Control’s (CDC) working groups on COVID-19 modeling and assuming that 5% of infectious people require a hospital bed, and 14% of hospitalized people require an ICU bed in each simulation (*33*). We then calculated the mean number of people-days (i.e., the number of people on a given day) where demand for hospitalization exceeds Utah’s capacity of 4,869 hospital beds and the number of people-days where demand for ICU beds exceeds Utah’s capacity of 687 ICU beds (*34, 35*). Note that these numbers are for total hospital and ICU beds, not those set aside for COVID-19 patients, and thus provide an upper bound for hospital capacity.

## Data Availability

All code is archived and freely available at doi:10.5281/zenodo.3924186.

doi:10.5281/zenodo.3924186

## List of Supplementary Materials

### Methods and materials

Clinical Prediction Rule Parametrization

### Supplementary figures

### Supplementary tables

## Acknowledgments

We thank the medical students who participated in the chart review process: Margaret Bale, Ben Berger. Jordan B. Peacock, William West, Alyssa Brown, Brendan Crabb, Sara Mann, and Valerie Martin.

## Author contributions

JRR, SMA, LTK, MJF, and DTL conceptualized this study. JRR, SMA, and RUS curated the data. JRR and SMA conducted the formal analyses. MJF and DTL acquired funding for this study. JRR, SMA, BB, MJF, and DTL developed the methodologic approach used in this study. JRR, SMA, BB, and LTK wrote the software for this study. LTK, MJF, and DTL supervised the work in this study. JRR developed all visualizations for this study. JRR and SMA wrote the first draft of this manuscript. All authors reviewed and edited this manuscript.

## Data and materials availability

All code is archived and freely available at doi:10.5281/zenodo.3924186.

## Supplementary materials and methods

### Clinical Prediction Rule

In additional to the CPR presented in the main text, we also explored three additional CPRs: A) a prediction rule using the same predictive variables as described previously in the main text, but NOT considering symptoms as potential predictors; B) a CPR using the same predictive variables as the main text AND continuous variables for SpO2 and pulse rate as collected by clinic personnel; C) a CPR using the same predictive variables as the main text AND a single variable for race/ethnicity, categorized as non-Hispanic White/Caucasian, Hispanic/Latino, Black or African American, Asian, Native Hawaiian and Other Pacific Islander, Other (including American Indian and Alaska Native), Choose not to disclose, or Unknown. Data necessary for the CPR presented in the main text can be collected from persons over the phone, before they present at a clinic/hospital, whereas supplementary Model B is a more complete set of data available at clinic presentation. Data for supplementary Model A is data automatically available from the electronic medical record along with contact tracing information.

### Parameterization

We first outline how we obtained our base set of parameter values, followed by an explanation of the range of each parameter that we considered in light of the uncertainty in these estimates.

To get estimates of testing demand in Utah, we assumed that Utah has had excess testing capacity since April 20, 2020, with a stated testing capacity of at least 6000 tests per day. Daily estimates of *R*_*t*_ for Utah were obtained for April 20 through May 20, 2020 (*36*). The average *R*_*t*_ value was 1.08, with no significant increase over that time (linear regression; t = -1.53, p > 0.1), suggesting that prevalence was approximately constant over that period. Using data from coronavirus.utah.gov (*21*), the mean total daily testing number over this period was 4207 tests with an average of 181 new positive tests each day. To consider the corresponding daily deaths resulting from cases over this period, we assumed a lag of 11 days (*37*), considering the daily deaths from May 1-June 1, 2020, with data collected from the Covid Tracking Project (*38*). Over this period, the average number of daily deaths was 2.09. We assumed an infection fatality rate (IFR) of 0.5%, suggesting an average of approximately 418 new cases per day over this period. Assuming a daily recovery probability of ∼1/6 (γ in the SEIR model), an infectious individual has a 5/6 probability of not recovering on a given day. Thus, at the start of a given day, the number of infected individuals is the sum of those becoming infectious that day (∼418 people), plus the number who became infectious the day before and did not yet recover (∼418*(5/6)), plus the number who became infectious two days before and have not yet recovered (∼418*(5/6)*(5/6)), etc. This is a geometric series which converges to an estimate of 2508, our daily estimate of the average prevalence over this three-week period. This implies that 7.2% (181/2508) of infectious people are test eligible daily, i.e., w_I_ = 0.072. Assuming the infectious period lasts for an average of 6 days, this implies that an individual has a ∼36% chance of getting tested during their time in the infectious class.

The average number of new negative tests each day was 4021. We assumed almost all of those came from susceptible individuals (rather than exposed or recovered), so that the proportion of individuals from the *S* class is approximately 4021/(total population of Utah). For a total population of approximately 3.2 million in Utah, this means *w*_*S*_ = 0.0013 (i.e., 4021/3200000). We assumed *w*_*E*_*= w*_*S*_, as those in the exposed class are typically pre-symptomatic and thus do not seek testing at a higher rate. In the absence of data, we assumed *w*_*R*_ = (1-0.36)\**w*_*S*_ = 0.00084, as ∼36% of recovered individuals should have been tested while infectious (based on the calculations above) and thus do not seeking testing again.

To explore how sensitive these outputs are to the IFR, we varied the IFR from 0.2-1%, as recommended by the CDCs working groups on modeling COVID-19. For all these values, *w*_*S*_ and *w*_*E*_ remained at 0.0013 and *w*_*R*_ decreased slightly, from *w*_*R*_ = 0.0011 to 0.00052. *w*_*I*_ had the highest sensitivity to the IFR, with *w*_*I*_ = 0.029, 0.072, 0.101, 0.144 for IFR = 0.2, 0.5, 0.7, and 1%, respectively. We considered *W*_*I*_ values ranging from 0.029 through 0.144 to determine the effect of this parameter on model outputs. We used a value of 0.7 for *<u>p</u>*, the proportion of exposed individuals who, if tested, would test positive for COVID-19. To test sensitivity to this value, we also considered *p =* 0.5 and 0.9.

## Supplementary figures

**Fig S1.**
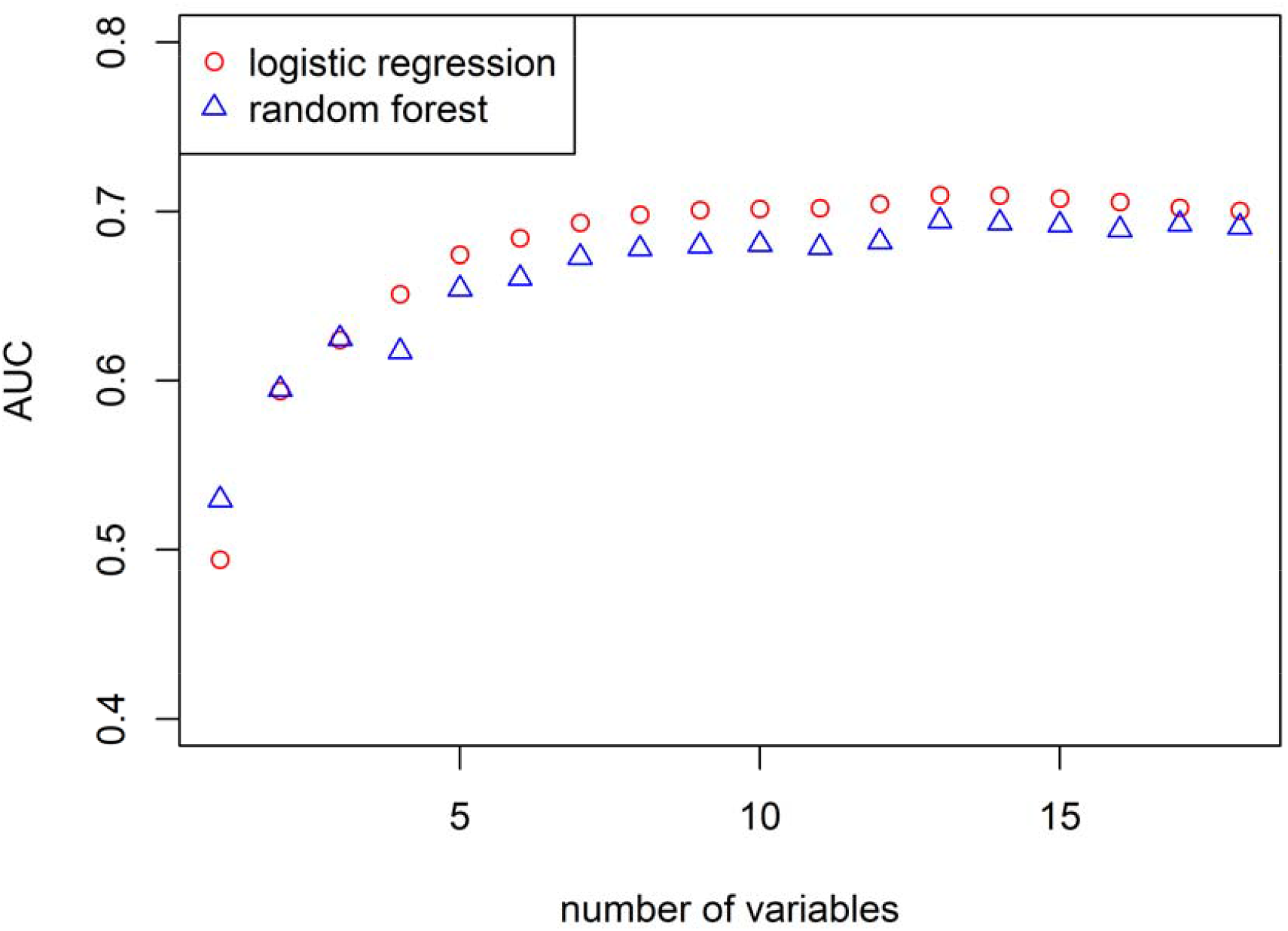
Comparison of AUC achieved by number of predictive variables included, and logistic versus random forest regression. All variables listed in the “main text model” of Table S1 were considered. The final CPR included in the main text included the top-5 predictors, as this balanced prediction with parsimony.

## Supplementary tables

**Table S1.**
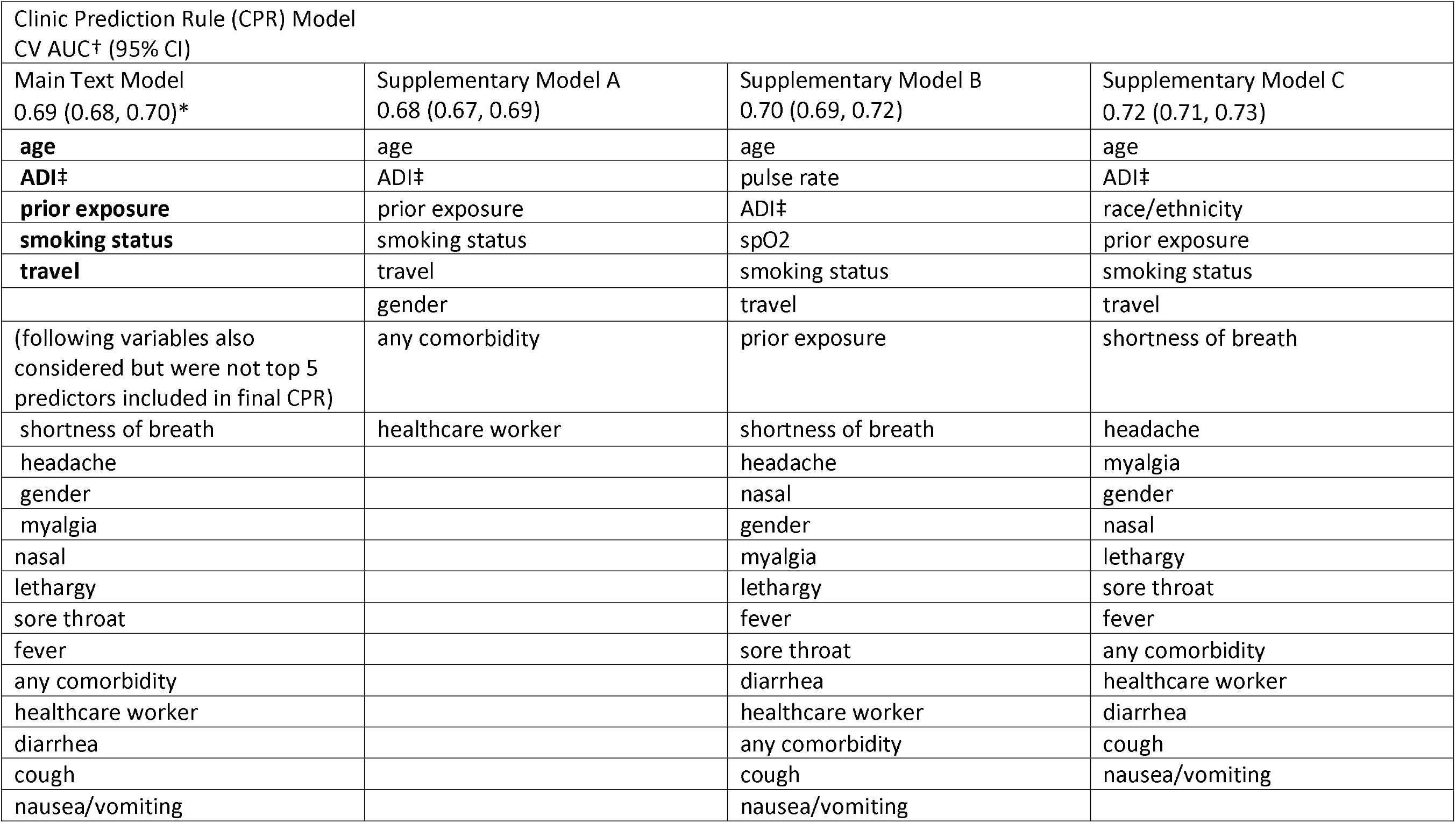
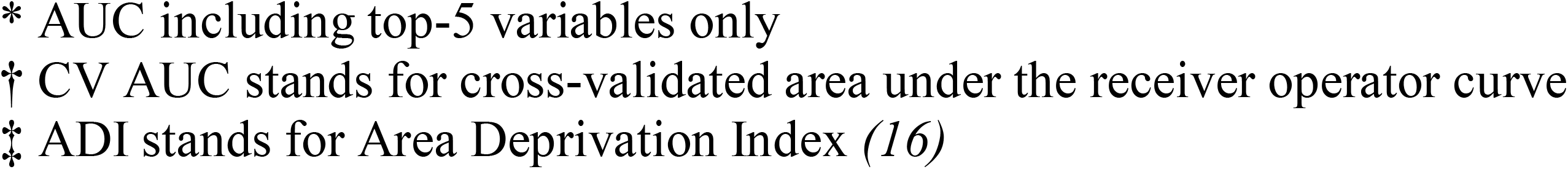
Clinical Prediction Rule (CPR) models explored and their cross-validated AUCs. All CPRs were estimated using logistic regression. All variables considered for each model are listed per column, ranked from most to least predictive. Bolded entries denote variables used in the final 5-variable CPR presented in the main text.

| Clinic Prediction Rule (CPR) Model<br>CV AUC† (95% CI) |  |  |  |
| --- | --- | --- | --- |
| Main Text Model<br>0.69 (0.68, 0.70)* | Supplementary Model A<br>0.68 (0.67, 0.69) | Supplementary Model B<br>0.70 (0.69, 0.72) | Supplementary Model C<br>0.72 (0.71, 0.73) |
| **age** | age | age | age |
| **ADI‡** | ADI‡ | pulse rate | ADI‡ |
| **prior exposure** | prior exposure | ADI‡ | race/ethnicity |
| **smoking status** | smoking status | spO2 | prior exposure |
| **travel** | travel | smoking status | smoking status |
|  | gender | travel | travel |
| (following variables also considered but were not top 5 predictors included in final CPR) | any comorbidity | prior exposure | shortness of breath |
| shortness of breath | healthcare worker | shortness of breath | headache |
| headache |  | headache | myalgia |
| gender |  | nasal | gender |
| myalgia |  | gender | nasal |
| nasal |  | myalgia | lethargy |
| lethargy |  | lethargy | sore throat |
| sore throat |  | fever | fever |
| fever |  | sore throat | any comorbidity |
| any comorbidity |  | diarrhea | healthcare worker |
| healthcare worker |  | healthcare worker | diarrhea |
| diarrhea |  | any comorbidity | cough |
| cough |  | cough | nausea/vomiting |
| nausea/vomiting |  | nausea/vomiting |  |

\* AUC including top-5 variables only
† CV AUC stands for cross-validated area under the receiver operator curve
‡ ADI stands for Area Deprivation Index (*16*)

**Table S2.** Effects of prioritized testing on the SEIR infection dynamics over additional clinical prediction rules and an additional range of parameter values. All parameter values are as stated in the text except as stated otherwise in this table. Each column compares the model with prioritized testing to one with indiscriminate testing. Bolded entries denote the results for the base parameter set described in the text, and are included for reference in each subsection.

|  | Delay in<br>peak timing<br>(days) | Reduction in<br>peak height<br>(people) (%) | Reduction in<br>total infections<br>(people) | Reduction in<br>people-days<br>above hospital<br>capacity (%) | Reduction in<br>people-days<br>above ICU<br>capacity (%) |
| --- | --- | --- | --- | --- | --- |
| **Model (AUC)** |  |  |  |  |  |
| **main model (0.69)** | **27** | **32,321 (31%)** | **40,933 (2%)** | **76,401 (85%)** | **108,707 (98%)** |
| A (0.68) | 30 | 29,207 (27%) | 40,138 (2%) | 71,981 (82%) | 108,354 (98%) |
| B (0.70) | 23 | 23,647 (21%) | 36,033 (2%) | 58,305 (68%) | 106,483 (97%) |
| C (0.72) | 23 | 34,933 (33%) | 35,045 (1%) | 84,509 (87%) | 111,478 (99%) |
| *p* |  |  |  |  |  |
| 0.5 | 32 | 29087 (27%) | 43,896 (2%) | 71,333 (80%) | 108,414 (98%) |
| **0.7** | **27** | **32,321 (31%)** | **40,933 (2%)** | **76,401 (85%)** | **108,707 (98%)** |
| 0.9 | 26 | 27,616 (26%) | 39,753 (2%) | 69,283 (82%) | 108,129 (98%) |

